# Does *COMT* Play a Role in Parkinson’s Disease Susceptibility Across Diverse Ancestral Populations?

**DOI:** 10.1101/2025.04.11.25325572

**Authors:** Miguel Martín-Bórnez, Nisar Shar, Mohamed Ahmed Nour, David Murphy, Inas Elsayed, Shri N Megha, Francisca Nwaokorie, Adedunni Olusanya, Nicole Kuznetsov, Sara Bandres-Ciga, Alastair J Noyce, Hirotaka Iwaki, Lietsel Jones, Pilar Gómez-Garre, Pablo Mir, the Global Parkinson’s Genetics Program (GP2), Maria Teresa Periñan

## Abstract

**Background:** The catechol-O-methyltransferase (*COMT*) gene is involved in brain catecholamine metabolism, but its association with Parkinson’s disease (PD) risk remains unclear.

**Objective:** To investigate the relationship between *COMT* genetic variants and PD risk across diverse ancestries.

**Methods:** We analyzed *COMT* variants in 2,251 PD patients and 2,835 controls of European descent using whole-genome sequencing from the Accelerating Medicines Partnership-Parkinson Disease (AMP-PD), along with 20,427 PD patients and 11,837 controls from 10 ancestries using genotyping data from the Global Parkinson’s Genetics Program (GP2).

**Results:** Utilizing the largest case-control datasets to date, no significant enrichment of *COMT* risk alleles in PD patients was observed across any ancestry group after correcting for multiple testing. Among Europeans, no correlations with cognitive decline, motor function, motor complications, or time to LID onset were observed.

**Conclusions:** These findings emphasize the need for larger, diverse cohorts to confirm the role of *COMT* in PD development and progression.

## Introduction

Parkinson’s disease (PD) is a progressive neurodegenerative disorder marked by motor symptoms such as tremors, rigidity, and bradykinesia, alongside diverse non-motor manifestations. Its etiology involves a complex interplay of genetic and environmental factors [1,2].

The enzyme catechol-O-methyltransferase (COMT; OMIM:116790) plays a crucial role in catecholamine metabolism in the brain, including dopamine and norepinephrine. *COMT* variants can affect enzyme activity and dopamine metabolism [3]. While numerous studies have explored associations between *COMT* variants and PD susceptibility [4], particularly the Val158Met (rs4680), and their impact on levodopa-induced dyskinesia (LID) [5,6], cognitive decline [7–9], and motor fluctuations [10], findings remain inconsistent, leaving the role of *COMT* and variants in close proximity unresolved. Noteworthy, *COMT* is located within the 1.5 Mb region deleted in 22q11.2 deletion syndrome, a genetic disorder caused by the deletion of a small chromosome 22 segment, typically resulting in a wide variety of symptoms, including neurological [11].

We comprehensively investigated whether genetic variation within *COMT* influences PD risk, LID, cognitive impairment, motor function, and complications, utilizing large-scale array-based genotyping and whole-genome sequencing (WGS) data from the Global Parkinson’s Genetics Program (GP2) [12] and the Accelerating Medicines Partnership - Parkinson Disease (AMP-PD) initiatives.

## Methods

We analyzed AMP-PD WGS data release 3.0 (https://amp-pd.org/), including 2,251 unrelated PD patients and 2,835 controls of European descent (Supplementary Table 1). Additionally, we utilized large-scale genotyping imputed data from GP2 release 7 (https://gp2.org/), comprising 20,427 PD patients and 11,837 controls from ten ancestry populations: European (EUR), African Admixed (AAC), African (AFR), Ashkenazi Jews (AJ), American Admixed (AMR), Central Asian (CAS), East Asian (EAS), Middle Eastern (MDE), South Asian (SAS), and Complex Admixture History (CAH).

The GenoTools pipeline (https://github.com/GP2code/GenoTools) was used for genetic ancestry prediction, and quality control (QC) on genotyping data was conducted according to previously described methods [13]. Variants with a Hardy-Weinberg Equilibrium (HWE) p-value≤1x10^-4^ in control samples were removed after preliminary QC. Variants were further pruned to exclude those with a minor allele frequency (MAF)≤1% and a minor allele count of 2. *COMT* gene positions were obtained from Ensembl (https://www.ensembl.org). Variants were annotated using ANNOVAR [14]. Gene-based burden analysis was performed to assess the cumulative effect of potentially functional (variants annotated as frameshift, nonframeshift, startloss, stoploss, stopgain, splicing, missense, exonic, untranslated region at the 5’ end (UTR5), untranslated region at the 3’ end (UTR3), upstream [-100bp], downstream [+100bp], or ncRNA), coding (variants annotated as frameshift, nonframeshift, startloss, stoploss, stopgain, splicing, or missense) and loss-of-function (variants annotated as frameshift, startloss, stopgain, or splicing) variants on PD risk using RVTESTS [15]. SNP-phenotype association analyses were performed with a generalized linear model in PLINK 2.0 [16]. In the logistic regression analysis, we included sex, age, and the first five genetic principal components (PCs) as covariates to account for population stratification. The Bonferroni correction was applied to adjust the p-values, accounting for all *COMT* variants within each ancestry independently. Power calculations were performed using the online Genetic Association Study (GAS) Power Calculator (https://csg.sph.umich.edu/abecasis/cats/gas_power_calculator/index.html).

We examined clinical data of individuals of European ancestry in GP2 and AMP-PD to explore the impact of *COMT* variants on cognitive and motor function, as well as LID development. Cognitive decline was assessed using MoCA scores (N for GP2=568; N for AMP-PD=1341) and linear regression models, adjusting for age, sex, education level, and PCs. We evaluated associations with MDS-UPDRS Parts III (N for GP2= 1268; N for AMP-PD=1666) and IV (N for GP2=359; N for AMP-PD=1794) using the same covariates. LID associations were assessed using Cox proportional hazards models, incorporating time-to-event data for LID onset and adjusting for relevant covariates (age, sex, PCs, and levodopa equivalent daily dose). Kaplan-Meier survival curves were used to visualize time-to-LID onset across *COMT* variants.

## Results

Leveraging the AMP-PD WGS data, we identified 491 variants within the *COMT* gene region, including 444 intronic, 13 exonic (8 synonymous, 4 missense, and 1 nonframeshift (3-bp) deletion), and 34 UTR variants (Supplementary Table 2). Association tests for PD risk revealed no significant results for any *COMT* variants (Supplementary Table 3).

For the GP2 genotyping imputed data, all identified variants are listed in Supplementary Table 2. In the GP2 CAS ancestry group, nominally significant associations with PD risk were observed for p.Val158Met (rs4680) (Effect Allele=A; Odds Ratio [OR]=0.690, 95% Confidence Interval [CI]=0.539-0.883, p=0.003, Bonferroni-corrected p=0.391) and p.His62= (rs4633) (Effect Allele = T; OR=0.678, 95% CI=0.528-0.870, p=0.002, Bonferroni-corrected p=0.280) (Figure 1, Supplementary Table 4). In the GP2 EUR ancestry group, p.Val158Met (Effect Allele=A; OR=0.932, 95% CI=0.878-0.989, p=0.020, Bonferroni-corrected p=1), p.His62= (Effect Allele=T; OR=0.935, 95% CI=0.881-0.992, p=0.026, Bonferroni-corrected p=1), and c.-98A>G (rs6269) (Effect Allele=G; OR=1.067, 95% CI=1.067-1.133, p=0.035, Bonferroni-corrected p=1) were nominally significantly associated with PD risk. Additionally, the c.*764C>T (rs165728) variant was nominally significantly associated with PD risk in the GP2 AJ (Effect Allele=C; OR=0.602, 95% CI=0.373-0.971, p=0.038, Bonferroni-corrected p=1) and MDE (Effect Allele=C; OR=2.475, 95% CI=1.03-5.949, p=0.043,

**Figure 1.**
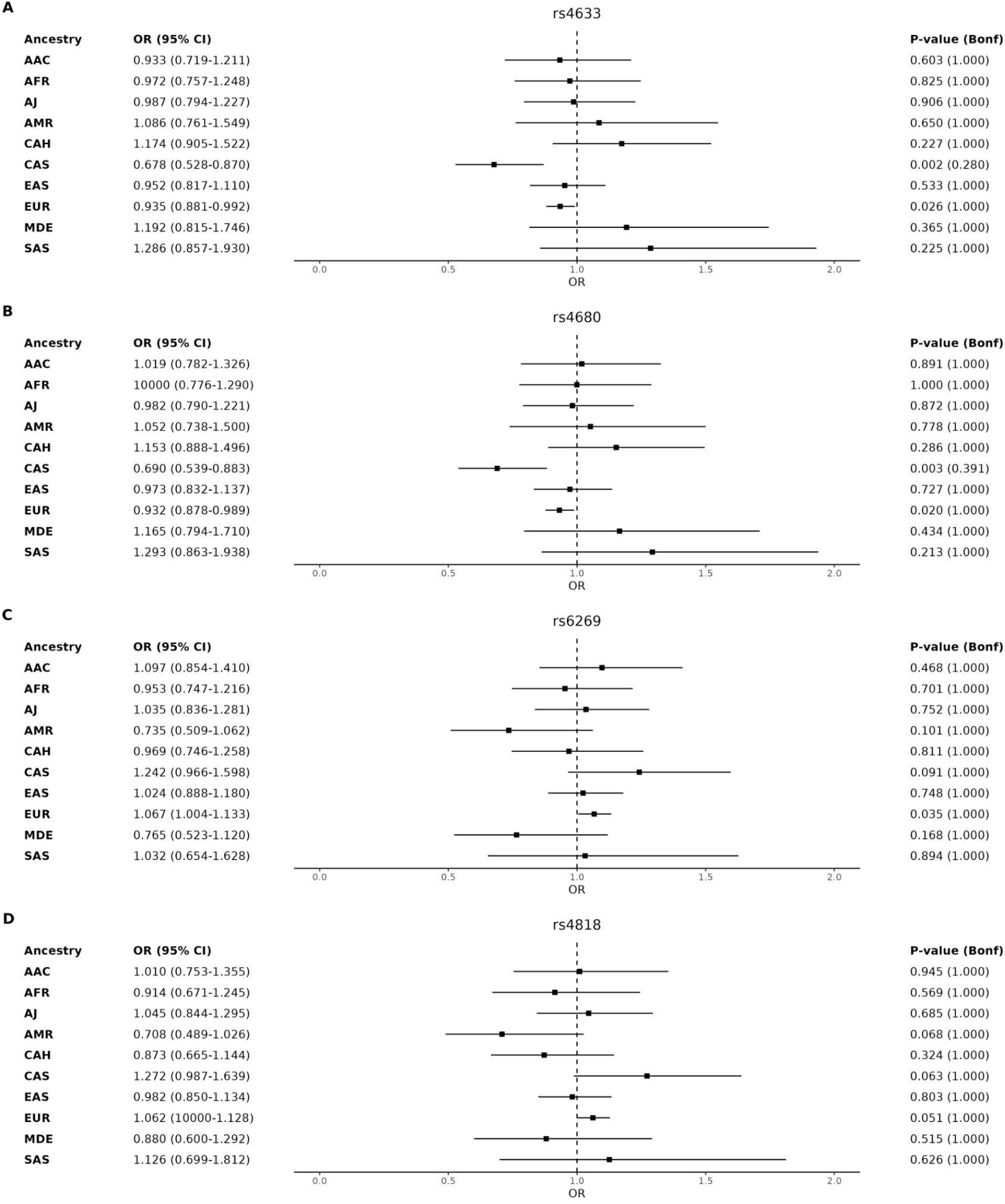
Forest plot illustrating the association between Parkinson’s disease risk and the four most extensively studied *COMT* variants (rs4633, rs4680, rs6269, rs4818) across ten ancestries. The analysis was performed using a generalized linear model adjusted for age, sex, and five principal components. **(A)** rs4633, **(B)** rs4680, **(C)** rs6269, and **(D)** rs4818. Abbreviations: AAC, African Admixed; AFR, African; AMR, American Admixed; Bonf, bonferroni; CAS, Central Asian; CI, confidence intervals; EAS, East Asian; EUR, European; MDE, Middle Eastern; OR, odds ratio; SAS, South Asian.

Bonferroni-corrected p=1) ancestry groups. Finally, p.Ala72Ser (rs6267) was found to be nominally significantly associated with PD risk in the EAS ancestry group (Effect Allele=T; OR=0.643, 95% CI=0.415-0.998, p=0.049, Bonferroni-corrected p=1). However, these associations did not remain significant after multiple testing corrections. Analysis of the latest PD risk GWAS summary statistics [17–21] did not identify significant associations (p-value threshold of 5×10^-8^) with *COMT* genetic variation (Supplementary Figure 1).

Gene-based burden analysis evaluated the cumulative effects of specific *COMT* variant sets on PD risk. In the GP2 AJ ancestry group, coding variants showed a nominal significant association with PD risk (N=3; SKAT p=0.069, SKAT-O p=0.036). Similarly, in the GP2 EUR ancestry group, coding variants (N=18; SKAT p=0.020, SKAT-O p=0.019) but also potentially functional variants were significantly associated with PD risk (N=105; SKAT p=0.032, SKAT-O p=0.037). However, none of these associations remained significant after multiple testing corrections and therefore these results should be interpreted with caution (Supplementary Table 5). In AMP-PD, potentially functional variants showed suggestive nominal evidence of association (N=46; SKAT p=0.071, SKAT-O p=0.110). Since loss-of-function variants were absent in all the groups examined, our study did not perform a burden analysis on them.

Regarding the role of *COMT* in influencing cognitive, motor function and complications in PD, both p.Val158Met (Effect Allele=G; BETA=-0.338; SE=0.150, p=0.024, Bonferroni-corrected p=1) and p.His62= (Effect Allele=C; BETA=-0.330; SE=0.149, p=0.027, Bonferroni-corrected p=1) showed nominal associations with MoCA scores in the GP2 EUR ancestry group. Similarly, these two variants were nominally associated with MDS-UPDRS Part IV scores in the AMP-PD WGS data, along with c.*764C>T (Effect Allele=T; BETA=0.666; SE=0.320, p=0.038, Bonferroni-corrected p=1). However, none of these associations remained significant after correction for multiple testing (Supplementary Tables 6–9).

The risk of developing LID was evaluated in 463 European PD patients from GP2, 34.3% of whom developed LID. The analysis focused on four *COMT* variants (rs6269, rs4633, rs4818, and rs4680) based on their functional significance, as they collectively modulate COMT activity [22]. No significant associations with time-to-LID onset were found in the Cox proportional hazards model (p-value>0.05). Kaplan-Meier survival curves are presented in Supplementary Figure 2.

## Discussion

This study leveraged AMP-PD WGS and GP2 genotyping data, representing the largest and most ancestrally diverse dataset to date, spanning ten different ancestries. It provides a unique and comprehensive analysis of *COMT* variants and their impact on PD risk across populations. Recent work by Poplawska et al. (2024) emphasizes the lack of ethnic diversity in clinical trials involving COMT inhibitors, underscoring the importance of our study in addressing the inclusion of underrepresented populations in research on *COMT* variants and PD risk [23].

No statistically significant associations were found for any ancestral populations in our analysis, including EAS, SAS and EUR, which have been the primary focus of previous research [4,9,24,25]. Notably, significant associations have been reported in Asian populations, particularly Japanese and Chinese cohorts (EAS), and to a lesser extent in Indian cohorts (SAS) [24–26]. A study involving 109 Japanese PD patients and 153 controls found a significant association with the homozygous p.Val158Met genotype over two decades ago [27]. This association was further supported by a meta-analysis of Asian populations, including 1,581 PD patients and 1,376 controls [26]. Despite having adequate statistical power (80% power to detect an OR≥1.5 for a MAF≥10%), our analysis of a large EAS cohort, consisting of 2,646 PD patients and 2,453 controls, did not replicate these findings. Similarly, in SAS populations, a meta-analysis of 489 Indian PD patients and 823 controls found a significant association between p.Val158Met and PD risk [25], but our study was unable to replicate this result. Future research should focus on increasing sample sizes in these populations to improve robustness.

Our study is the largest to date investigating p.Val158Met in European ancestry. A previous meta-analysis of 9,719 PD patients and 14,634 controls found no significant association between this variant and PD risk in European populations [4]. Another meta-analysis of 11,428 PD cases and 16,726 controls also reported no significant associations for p.Val158Met in either European or Asian ancestries [9]. Our results are consistent with these findings, further supporting the lack of association with sufficient statistical power.

Levodopa-induced dyskinesia (LID) is a major complication of long-term levodopa therapy in PD, affecting 20%-40% of patients [28]. In our study, 34.3% of patients developed LID, all of whom were on levodopa treatment. While we found no significant associations between *COMT* variants and LID in European ancestry, our findings align with a recent meta-analysis, which also reported no genome-wide significant associations between *COMT* variants and time-to-LID [28]. However, previous studies have suggested that the rs4680 genotype may modulate LID risk in PD patients from European [29], Asian and Brazilian ancestries [5,6,10].

We also examined the relationship between *COMT* variants and motor function, as assessed by MDS-UPDRS Parts III (motor function) and IV (motor complications, including LID). We found no significant associations with motor function or complications in the GP2 EUR ancestry group after correction for multiple comparisons. A meta-analysis of 1,574 Asian PD patients reported a significant association between rs4680 and higher UPDRS III scores [9], but we did not replicate this in Europeans. This may reflect population-specific genetic architecture or limited power for smaller effect sizes.

Similarly, we found no significant association with *COMT* variants and cognitive decline in the GP2 EUR ancestry group. This aligns with results from a meta-analysis of p.Val158Met, which also reported no significant link to cognitive decline in PD [9]. However, a longitudinal study involving 246 PD patients suggested that homozygous carriers of rs4680 may experience accelerated cognitive decline [30].

In conclusion, our results show no association between *COMT* and PD risk that passes multiple test correction, emphasizing the need for larger, diverse cohorts to confirm its role in PD development and progression.

## Data Availability

Data used in the preparation of this article were obtained from the Global Parkinson′s Genetics Program (GP2; https://gp2.org). Specifically, we used Tier 2 data from GP2 release 7 (DOI:10.5281/zenodo.10962119).
All code generated for this article, and the identifiers for all software programs and packages used, are available on GitHub [https://github.com/GP2code/COMT-PD-GeneAnalysis] and were given a persistent identifier via Zenodo [DOI 10.5281/zenodo.15185052].

## Competing interests

The authors report no competing interests.

## Data Sharing

Data used in the preparation of this article were obtained from the Global Parkinson’s Genetics Program (GP2; https://gp2.org). Specifically, we used Tier 2 data from GP2 release 7 (DOI:10.5281/zenodo.10962119). Tier 1 data can be accessed by completing a form on the Accelerating Medicines Partnership in Parkinson’s Disease (AMP®-PD) website (https://amp-pd.org/register-for-amp-pd). Tier 2 data access requires approval and a Data Use Agreement signed by your institution. All code generated for this article, and the identifiers for all software programs and packages used, are available on GitHub [https://github.com/GP2code/COMT-PD-GeneAnalysis] and were given a persistent identifier via Zenodo [DOI 10.5281/zenodo.15185052].

## Acknowledgments

This work was carried out with the support and guidance of the ‘GP2 Trainee Network’ which is part of the Global Parkinson’s Genetics Program and funded by the Aligning Science Across Parkinson’s (ASAP) initiative. Data used in the preparation of this article were obtained from Global Parkinson’s Genetics Program (GP2). For a complete list of GP2 members, see https://gp2.org. Data used in the preparation of this article were obtained from the Accelerating Medicines Partnership^®^ (AMP^®^) Parkinson’s Disease (AMP^®^ PD) Knowledge Platform. For up-to-date information on the study, visit https://www.amp-pd.org. ACCELERATING MEDICINES PARTNERSHIP and AMP are registered service marks of the US Department of Health and Human Services.

## Funding

This research was supported in part by the Intramural Research Program of the NIH, National Institute on Aging (NIA), National Institutes of Health, Department of Health and Human Services; project number ZO1 AG000535 and ZIA AG000949, as well as the National Institute of Neurological Disorders and Stroke (NINDS) and the National Human Genome Research Institute (NHGRI). This project was supported by the Global Parkinson’s Genetics Program (GP2; https://gp2.org). GP2 is funded by the Aligning Science Across Parkinson’s (ASAP) initiative and implemented by The Michael J. Fox Foundation for Parkinson’s Research (MJFF). For a complete list of GP2 members see doi.org/10.5281/zenodo.7904831. The AMP^®^ PD program is a public-private partnership managed by the Foundation for the National Institutes of Health and funded by the National Institute of Neurological Disorders and Stroke (NINDS) in partnership with the Aligning Science Across Parkinson’s (ASAP) initiative; Celgene Corporation, a subsidiary of Bristol-Myers Squibb Company; GlaxoSmithKline plc (GSK); The Michael J. Fox Foundation for Parkinson’s Research; Pfizer Inc.; Sanofi US Services Inc.; and Verily Life Sciences. Miguel Martín-Bórnez is supported by a predoctoral contract for training in health research (PFIS) from the Instituto de Salud Carlos III (FI22/00226). Megha Shri N received funding support from CSIR under sanction no. 09/0490(16118)/2022-EMR-I and the Parkinson’s Disease and Movement Disorders Research Fund (File no. 13020).

## Author contributions

M.T.P conceptualized the manuscript. M.M-B, N.S., M.A.N., D.M., I.E., M.S.N., F.N., A.O., and M.T.P wrote the first manuscript draft. All authors reviewed, edited, and approved the final version of the manuscript for submission.

## Notes

### Competing Interest Statement

The authors have declared no competing interest.

### Author Declarations

Data used in the preparation of this article were obtained from the Global Parkinson′s Genetics Program (GP2; https://gp2.org). Specifically, we used Tier 2 data from GP2 release 7 (DOI:10.5281/zenodo.10962119). Tier 1 data can be accessed by completing a form on the Accelerating Medicines Partnership in Parkinson′s Disease (AMP®-PD) website (https://amp-pd.org/register-for-amp-pd). Tier 2 data access requires approval and a Data Use Agreement signed by your institution.

## References

1. Bloem BR, Okun MS, Klein C. Parkinson’s disease. Lancet. 2021;397:2284–303.

2. Bandres-Ciga S, Diez-Fairen M, Kim JJ, Singleton AB. Genetics of Parkinson’s disease: An introspection of its journey towards precision medicine. Neurobiol Dis. 2020;137:104782.

3. Chen J, Lipska BK, Halim N, Ma QD, Matsumoto M, Melhem S, et al. Functional analysis of genetic variation in catechol-O-methyltransferase (COMT): effects on mRNA, protein, and enzyme activity in postmortem human brain. Am J Hum Genet. 2004;75:807–21.

4. Jiménez-Jiménez FJ, Alonso-Navarro H, García-Martín E, Agúndez JAG. COMT gene and risk for Parkinson’s disease: a systematic review and meta-analysis. Pharmacogenet Genomics. 2014;24:331–9.

5. Dwivedi A, Dwivedi N, Kumar A, Singh VK, Pathak A, Chaurasia RN, et al. Association of Catechol-O-Methyltransferase Gene rs4680 Polymorphism and Levodopa Induced Dyskinesia in Parkinson’s Disease: A Meta-Analysis and Systematic Review. J Geriatr Psychiatry Neurol. 2023;36:98–106.

6. Yin Y, Liu Y, Xu M, Zhang X, Li C. Association of COMT rs4680 and MAO-B rs1799836 polymorphisms with levodopa-induced dyskinesia in Parkinson’s disease-a meta-analysis. Neurol Sci. 2021;42:4085–94.

7. Barnett JH, Scoriels L, Munafò MR. Meta-analysis of the cognitive effects of the catechol-O-methyltransferase gene Val158/108Met polymorphism. Biol Psychiatry. 2008;64:137–44.

8. Lin C-H, Fan J-Y, Lin H-I, Chang C-W, Wu Y-R. Catechol-O-methyltransferase (COMT) genetic variants are associated with cognitive decline in patients with Parkinson’s disease. Parkinsonism Relat Disord. 2018;50:48–53.

9. Tang C, Wang W, Shi M, Zhang N, Zhou X, Li X, et al. Meta-Analysis of the Effects of the Catechol-O-Methyltransferase Val158/108Met Polymorphism on Parkinson’s Disease Susceptibility and Cognitive Dysfunction. Front Genet. 2019;10:454351.

10. Soraya GV, Ulhaq ZS, Shodry S, A’raaf Sirojan Kusuma M, Herawangsa S, Sativa MO, et al. Polymorphisms of the dopamine metabolic and signaling pathways are associated with susceptibility to motor levodopa-induced complications (MLIC) in Parkinson’s disease: a systematic review and meta-analysis. Neurol Sci. 2022;43:3649–70.

11. Mok KY, Sheerin U, Simón-Sánchez J, Salaka A, Chester L, Escott-Price V, et al. Deletions at 22q11.2 in idiopathic Parkinson’s disease: a combined analysis of genome-wide association data. Lancet Neurol. 2016;15:585–96.

12. Global Parkinson’s Genetics Program. GP2: The global Parkinson’s genetics program. Mov Disord. 2021;36:842–51.

13. Vitale D, Koretsky MJ, Kuznetsov N, Hong S, Martin J, James M, et al. GenoTools: An Open-Source Python Package for Efficient Genotype Data Quality Control and Analysis. G3 (Bethesda) [Internet]. 2024; Available from: 10.1093/g3journal/jkae268

14. Wang K, Li M, Hakonarson H. ANNOVAR: functional annotation of genetic variants from high-throughput sequencing data. Nucleic Acids Res. 2010;38:e164.

15. Zhan X, Hu Y, Li B, Abecasis GR, Liu DJ. RVTESTS: an efficient and comprehensive tool for rare variant association analysis using sequence data. Bioinformatics. 2016;32:1423–6.

16. Hill A, Loh P-R, Bharadwaj RB, Pons P, Shang J, Guinan E, et al. Stepwise Distributed Open Innovation Contests for Software Development: Acceleration of Genome-Wide Association Analysis. Gigascience. 2017;6:1–10.

17. Loesch DP, Horimoto ARVR, Heilbron K, Sarihan EI, Inca-Martinez M, Mason E, et al. Characterizing the genetic architecture of Parkinson’s disease in Latinos. Ann Neurol. 2021;90:353–65.

18. Nalls MA, Blauwendraat C, Vallerga CL, Heilbron K, Bandres-Ciga S, Chang D, et al. Identification of novel risk loci, causal insights, and heritable risk for Parkinson’s disease: a meta-analysis of genome-wide association studies. Lancet Neurol. 2019;18:1091–102.

19. Foo JN, Chew EGY, Chung SJ, Peng R, Blauwendraat C, Nalls MA, et al. Identification of Risk Loci for Parkinson Disease in Asians and Comparison of Risk Between Asians and Europeans: A Genome-Wide Association Study. JAMA Neurol. 2020;77:746–54.

20. Kim JJ, Vitale D, Otani DV, Lian MM, Heilbron K, 23andMe Research Team, et al. Multi-ancestry genome-wide association meta-analysis of Parkinson’s disease. Nat Genet. 2024;56:27–36.

21. Rizig M, Bandres-Ciga S, Makarious MB, Ojo OO, Crea PW, Abiodun OV, et al. Identification of genetic risk loci and causal insights associated with Parkinson’s disease in African and African admixed populations: a genome-wide association study. Lancet Neurol. 2023;22:1015–25.

22. Nackley AG, Shabalina SA, Tchivileva IE, Satterfield K, Korchynskyi O, Makarov SS, et al. Human catechol-O-methyltransferase haplotypes modulate protein expression by altering mRNA secondary structure. Science. 2006;314:1930–3.

23. Schumacher-Schuh AF, Bieger A, Okunoye O, Mok KY, Lim S-Y, Bardien S, et al. Underrepresented populations in Parkinson’s genetics research: Current landscape and future directions. Mov Disord. 2022;37:1593–604.

24. Lechun L, Yu S, Pengling H, Changqi H. The COMT Val158Met polymorphism as an associated risk factor for Parkinson’s disease in Asian rather than Caucasian populations. Neurol India. 2013;61:12–6.

25. Wang Y-C, Zou Y-B, Xiao J, Pan C, Jiang S, Zheng Z-J, et al. COMT Val158Met polymorphism and Parkinson’s disease risk: a pooled analysis in different populations. Neurol Res. 2019;41:319–25.

26. Chuan L, Gao J, Lei Y, Wang R, Lu L, Zhang X. Val158Met polymorphism of COMT gene and Parkinson’s disease risk in Asians. Neurol Sci. 2015;36:109–15.

27. Kunugi H, Nanko S, Ueki A, Otsuka E, Hattori M, Hoda F, et al. High and low activity alleles of catechol-O-methyltransferase gene: ethnic difference and possible association with Parkinson’s disease. Neurosci Lett. 1997;221:202–4.

28. Martinez-Carrasco A, Real R, Lawton M, Iwaki H, Tan MMX, Wu L, et al. Genetic meta-analysis of levodopa induced dyskinesia in Parkinson’s disease. NPJ Parkinsons Dis. 2023;9:128.

29. de Lau LML, Verbaan D, Marinus J, Heutink P, van Hilten JJ. Catechol-O-methyltransferase Val158Met and the risk of dyskinesias in Parkinson’s disease. Mov Disord. 2012;27:132–5.

30. Paul KC, Rausch R, Creek MM, Sinsheimer JS, Bronstein JM, Bordelon Y, et al. APOE, MAPT, and COMT and Parkinson’s disease susceptibility and cognitive symptom progression. J Parkinsons Dis. 2016;6:349–59.

