## Supplementary Material for "Does *COMT* Play a Role in Parkinson’s Disease Susceptibility Across Diverse Ancestral Populations?"

**Supplementary Table 1.** Summary statistics of the GP2 genotyping and AMP-PD WGS.

*Abbreviations: AAC, African Admixed; AFR, African; AMR, American Admixed; CAS, Central Asian; SD, Standard deviation; EAS, East Asian; EUR, European; MDE, Middle Eastern; SAS, South Asian; CAH, Complex Admixture History; WGS, Whole Genome Sequencing.*

| **Ancestry** | **Cohort** | ***N* Total**  ***(n* PD*, n* controls*)*** | **Age, *y* (Mean ± SD)** | **Male sex, *n* (%)** |
| --- | --- | --- | --- | --- |
| AAC | GP2 | 1072 (279, 793) | 65.83 ± 10.37 | 437 (40.8) |
| AFR | GP2 | 2546 (929, 1617) | 63.48 ± 14.92 | 1432 (56.2) |
| AJ | GP2 | 1625 (1234, 391) | 69.76 ± 9.85 | 1097 (67.5) |
| AMR | GP2 | 577 (427,150) | 61.83 ± 11.95 | 327 (56.7) |
| CAH | GP2 | 777 (494, 283) | 55.28 ± 17.42 | 409 (52.6) |
| CAS | GP2 | 825 (519, 306) | 59.03 ± 9.64 | 386 (46.8) |
| EAS | GP2 | 4892 (2547, 2345) | 64.96 ± 10.96 | 3105 (63.5) |
| SAS | GP2 | 517 (319, 198) | 59.50 ± 15.62 | 336 (65.0) |
| MDE | GP2 | 426 (230, 196) | 60.12 ± 11.28 | 267 (62.7) |
| EUR | GP2 | 18225 (13034, 5191) | 66.79 ± 11.21 | 11180 (61.3) |
|  | AMP-PD | 5086 (2251, 2835) | 66.91 ± 11.84 | 2803 (55.1) |

**Supplementary Table 2.** *COMT* variants identified in AMP-PD WGS and GP2 genotyping data.

*Abbreviations: AAC, African Admixed; AFR, African; AMR, American Admixed; CAS, Central Asian; CI, confidence intervals; EAS, East Asian; EUR, European; MDE, Middle Eastern; SAS, South Asian; CAH, Complex Admixture History; WGS, Whole Genome Sequencing.*

| **Cohort** | **Ancestry** | **Total Variants** | **Intronic** | **3’-UTR** | **5’-UTR** | **Coding** | | |
| --- | --- | --- | --- | --- | --- | --- | --- | --- |
|  |  |  |  |  |  | **Synonymous** | **Nonsynonymous** | **Nonframeshift deletion** |
| AMP-PD | EUR | 491 | 444 | 20 | 14 | 8 | 4 | 1 |
| GP2 | EUR | 971 | 865 | 39 | 32 | 15 | 18 | 2 |
|  | AFR | 476 | 432 | 21 | 9 | 6 | 8 | 0 |
|  | EAS | 412 | 366 | 15 | 12 | 11 | 8 | 0 |
|  | AMR | 245 | 219 | 12 | 5 | 7 | 2 | 0 |
|  | CAS | 224 | 207 | 6 | 4 | 5 | 2 | 0 |
|  | AJ | 242 | 214 | 12 | 6 | 7 | 3 | 0 |
|  | SAS | 200 | 180 | 8 | 3 | 5 | 4 | 0 |
|  | MDE | 200 | 181 | 8 | 3 | 6 | 2 | 0 |
|  | AAC | 406 | 372 | 14 | 11 | 5 | 4 | 0 |
|  | CAH | 357 | 324 | 15 | 8 | 6 | 4 | 0 |

**Supplementary Table 3.** *COMT exonic* variants associated with PD risk adjusted by age at baseline, sex, and five principal components in AMP-PD WGS data.

*Abbreviations: A1, Allele 1, effect allele; OR, odds ratio; CI, confidence intervals; BONF, Bonferroni p-value correction.*

| **Variant** | **Hom Cases** | **Het Cases** | **Total Cases** | **Carrier freq in Cases** | **Hom Controls** | **Het Controls** | **Total Controls** | **Carrier freq in Controls** | **A1** | **OR (95% CI)** | ***p* (BONF)** |
| --- | --- | --- | --- | --- | --- | --- | --- | --- | --- | --- | --- |
| chr22:19962429:A:G  (rs6269; c.-98A>G) | 415 | 1081 | 2251 | 0.664594 | 468 | 1377 | 2835 | 0.650794 | G | 1.098 (1.008 - 1.195) | 0.032 (1) |
| chr22:19962712:C:T  (rs4633; p.His62=) | 553 | 1116 | 2251 | 0.494 | 746 | 1424 | 2835 | 0.514 | T | 0.905 (0.832 - 0.984) | 0.019 (1) |
| chr22:19963684:C:G  (rs4818; p.Leu136=) | 411 | 1072 | 2251 | 0.421 | 453 | 1362 | 2835 | 0.400 | G | 1.109 (1.018 - 1.207) | 0.018 (1) |
| chr22:19963748:G:A  (rs4680; p.Val158Met) | 552 | 1114 | 2251 | 0.493 | 739 | 1426 | 2835 | 0.512 | A | 0.909 (0.836 - 0.989) | 0.027 (1) |
| chr22:19964281:G:A  (rs769224; p.Pro199=) | 1 | 65 | 2251 | 0.015 | 1 | 116 | 2835 | 0.021 | A | 0.718 (0.519 - 0.994) | 0.046 (1) |
| chr22:19964293:C:T  (rs165631; p.Leu203=) | 0 | 47 | 2251 | 0.010 | 3 | 101 | 2835 | 0.019 | T | 0.738 (0.508 - 1.071) | 0.109 (1) |
| chr22:19969030:C:G  (rs9332381; c.*294C>G) | 2 | 138 | 2251 | 0.032 | 4 | 205 | 2835 | 0.038 | G | 0.843 (0.667 - 1.066) | 0.153 (1) |
| chr22:19969258:G:A  (rs165599; c.*522G>A) | 194 | 941 | 2251 | 0.50422 | 261 | 1188 | 2835 | 0.511111 | A | 0.999 (0.912 - 1.096) | 0.991 (1) |
| chr22:19969340:G:A  (rs36082074; c.*604G>A) | 2 | 136 | 2251 | 0.031 | 3 | 159 | 2835 | 0.029 | A | 1.221 (0.954 - 1.563) | 0.113 (1) |
| chr22:19969362:T:C  (rs35478083; c.*626T>C) | 2 | 133 | 2251 | 0.030 | 3 | 201 | 2835 | 0.037 | C | 0.832 (0.656 - 1.055) | 0.130 (1) |
| chr22:19969500:C:T  (rs165728; c.*764C>T) | 5 | 213 | 2251 | 0.050 | 7 | 249 | 2835 | 0.046 | C | 0.995 (0.818 - 1.208) | 0.956 (1) |

**Supplementary Table 4.** *COMT exonic* variants associated with PD risk adjusted by age at baseline, sex, and five principal components in GP2 genotyping data.

*Abbreviations: A1, Allele 1, effect allele; OR, odds ratio; CI, confidence intervals; BONF, Bonferroni p-value correction; AAC: African Admixed; AFR: African; AJ: Ashkenazi Jews; AMR: American Admixed; CAS: Central Asian; EAS: East Asian; EUR: European; MDE: Middle Eastern; SAS: South Asian.*

| **GP2 Ancestry** | **Variant** | **Hom Cases** | **Het Cases** | **Total Cases** | **Carrier freq in Cases** | **Hom Controls** | **Het Controls** | **Total Controls** | **Carrier freq in Controls** | **A1** | **OR (95% CI)** | ***p* (BONF)** |
| --- | --- | --- | --- | --- | --- | --- | --- | --- | --- | --- | --- | --- |
| EUR | chr22:19962429:A:G  (rs6269; c.-98A>G) | 2221 | 6247 | 13034 | 0.416 | 807 | 2516 | 5191 | 0.404 | G | 1.067 (1.004-1.133) | 0.035 (1) |
| AFR |  | 135 | 392 | 929 | 0.357 | 230 | 747 | 1617 | 0.374 | G | 0.953 (0.747-1.216) | 0.700 (1) |
| EAS |  | 280 | 1110 | 2547 | 0.328 | 224 | 1042 | 2345 | 0.318 | G | 1.024 (0.888-1.18) | 0.748 (1) |
| AAC |  | 43 | 137 | 279 | 0.400 | 110 | 373 | 793 | 0.374 | G | 1.097 (0.854-1.410) | 0.468 (1) |
| AJ |  | 279 | 604 | 1234 | 0.471 | 87 | 194 | 391 | 0.471 | G | 1.035 (0.836-1.281) | 0.752 (1) |
| AMR |  | 48 | 176 | 427 | 0.319 | 20 | 76 | 150 | 0.387 | G | 0.735 (0.509-1.062) | 0.101 (1) |
| SAS |  | 38 | 128 | 319 | 0.320 | 18 | 88 | 198 | 0.313 | G | 1.032 (0.654-1.628) | 0.894 (1) |
| CAH |  | 66 | 224 | 494 | 0.361 | 33 | 142 | 283 | 0.369 | G | 0.969 (0.746-1.258) | 0.811 (1) |
| CAS |  | 77 | 269 | 519 | 0.408 | 48 | 128 | 306 | 0.366 | G | 1.242 (0.966-1.598) | 0.091 (1) |
| MDE |  | 43 | 100 | 230 | 0.404 | 57 | 87 | 196 | 0.513 | G | 0.765 (0.523-1.12) | 0.168 (1) |
| EUR | chr22:19962712:C:T  (rs4633; p.His62=) | 3152 | 6312 | 13034 | 0.484 | 1198 | 2506 | 5191 | 0.472 | T | 0.935 (0.881-0.992) | 0.026 (1) |
| AFR |  | 78 | 341 | 929 | 0.271 | 153 | 670 | 1617 | 0.306 | T | 0.972 (0.757-1.248) | 0.825 (1) |
| EAS |  | 155 | 795 | 2547 | 0.249 | 141 | 797 | 2345 | 0.266 | T | 0.952 (0.817-1.11) | 0.533 (1) |
| AAC |  | 23 | 140 | 279 | 0.333 | 96 | 359 | 793 | 0.347 | T | 0.933 (0.719-1.211) | 0.603 (1) |
| AJ |  | 263 | 625 | 1234 | 0.466 | 84 | 192 | 391 | 0.462 | T | 0.987 (0.794-1.227) | 0.906 (1) |
| AMR |  | 68 | 219 | 427 | 0.419 | 23 | 79 | 150 | 0.419 | T | 1.086 (0.761-1.549) | 0.65 (1) |
| SAS |  | 65 | 145 | 319 | 0.431 | 38 | 104 | 198 | 0.457 | T | 1.286 (0.857-1.93) | 0.225 (1) |
| CAH |  | 82 | 250 | 494 | 0.419 | 40 | 136 | 283 | 0.382 | T | 1.174 (0.905-1.522) | 0.227 (1) |
| CAS |  | 71 | 243 | 519 | 0.377 | 75 | 135 | 306 | 0.473 | T | 0.678 (0.528-0.87) | 0.002 (0.280) |
| MDE |  | 56 | 109 | 230 | 0.483 | 41 | 83 | 196 | 0.425 | T | 1.193 (0.815-1.746) | 0.365 (1) |
| EUR | chr22:19963684:C:G  (rs4818; p.Leu136=) | 2164 | 6224 | 13034 | 0.410 | 787 | 2499 | 5191 | 0.399 | G | 1.062 (1-1.128) | 0.051 (1) |
| AFR |  | 23 | 252 | 929 | 0.162 | 56 | 465 | 1617 | 0.180 | G | 0.914 (0.671-1.245) | 0.569 (1) |
| EAS |  | 268 | 1084 | 2547 | 0.325 | 206 | 1030 | 2345 | 0.314 | G | 0.982 (0.85-1.134) | 0.803 (1) |
| AAC |  | 12 | 108 | 279 | 0.238 | 39 | 262 | 793 | 0.215 | G | 1.010 (0.753-1.355) | 0.945 (1) |
| AJ |  | 273 | 610 | 1234 | 0.468 | 85 | 195 | 391 | 0.467 | G | 1.045 (0.844-1.295) | 0.685 (1) |
| AMR |  | 42 | 172 | 427 | 0.300 | 20 | 73 | 150 | 0.377 | G | 0.708 (0.489-1.026) | 0.068 (1) |
| SAS |  | 31 | 131 | 319 | 0.305 | 16 | 85 | 198 | 0.298 | G | 1.126 (0.699-1.812) | 0.626 (1) |
| CAS |  | 75 | 265 | 519 | 0.401 | 45 | 124 | 306 | 0.351 | G | 1.272 (0.987-1.639) | 0.063 (1) |
| CAH |  | 48 | 205 | 494 | 0.307 | 26 | 132 | 283 | 0.332 | G | 0.873 (0.665-1.144) | 0.324 (1) |
| MDE |  | 40 | 95 | 230 | 0.483 | 49 | 83 | 196 | 0.425 | G | 0.88 (0.6-1.292) | 0.515 (1) |
| EUR | chr22:19963748:G:A  (rs4680; p.Val158Met) | 3145 | 6356 | 13034 | 0.500 | 1331 | 2502 | 5191 | 0.513 | A | 0.932 (0.878-0.989) | 0.020 (1) |
| AFR |  | 71 | 320 | 929 | 0.249 | 117 | 653 | 1617 | 0.275 | A | 1.000 (0.776-1.290) | 1.000 (1) |
| EAS |  | 147 | 796 | 2547 | 0.248 | 131 | 787 | 2345 | 0.262 | A | 0.973 (0.832-1.137) | 0.727 (1) |
| AAC |  | 19 | 142 | 279 | 0.323 | 83 | 347 | 793 | 0.323 | A | 1.019 (0.782-1.326) | 0.891 (1) |
| AJ |  | 249 | 625 | 1234 | 0.455 | 76 | 196 | 391 | 0.445 | A | 0.982 (0.79-1.221) | 0.872 (1) |
| AMR |  | 71 | 217 | 427 | 0.420 | 23 | 79 | 150 | 0.419 | A | 1.052 (0.738-1.5) | 0.778 (1) |
| SAS |  | 63 | 140 | 319 | 0.420 | 37 | 98 | 198 | 0.434 | A | 1.293 (0.863-1.938) | 0.213 (1) |
| CAH |  | 79 | 243 | 494 | 0.407 | 38 | 133 | 283 | 0.369 | A | 1.153 (0.888-1.496) | 0.286 (1) |
| CAS |  | 72 | 248 | 519 | 0.378 | 76 | 135 | 306 | 0.472 | A | 0.69 (0.539-0.883) | 0.003 (0.391) |
| MDE |  | 57 | 107 | 230 | 0.485 | 40 | 86 | 196 | 0.476 | A | 1.165 (0.794-1.71) | 0.434 (1) |
| EUR | chr22:19964281:G:A  (rs769224; p.Pro199=) | 5 | 516 | 13034 | 0.020 | 7 | 231 | 5191 | 0.024 | A | 0.854 (0.704-1.036) | 0.108 (1) |
| AFR |  | 8 | 142 | 929 | 0.085 | 17 | 274 | 1617 | 0.095 | A | 0.855 (0.547-1.336) | 0.492 (1) |
| EAS |  | 15 | 316 | 2547 | 0.069 | 7 | 223 | 2345 | 0.051 | A | 0.828 (0.63-1.09) | 0.179 (1) |
| AAC |  | 1 | 35 | 279 | 0.066 | 7 | 131 | 793 | 0.091 | A | 0.862 (0.537-1.382) | 0.537 (1) |
| AJ |  | 0 | 47 | 1234 | 0.019 | 0 | 13 | 391 | 0.017 | A | 0.869 (0.396-1.905) | 0.726 (1) |
| AMR |  | 1 | 23 | 427 | 0.029 | 0 | 8 | 150 | 0.027 | A | 3.19 (0.724-14.059) | 0.125 (1) |
| CAH |  | 1 | 41 | 494 | 0.044 | 1 | 15 | 283 | 0.030 | A | 1.161 (0.599-2.249) | 0.659 (1) |
| SAS |  | 1 | 11 | 319 | 0.020 | 0 | 4 | 198 | 0.010 | A | 1.754 (0.403-7.642) | 0.454 (1) |
| CAS |  | 1 | 33 | 519 | 0.034 | 0 | 25 | 306 | 0.041 | A | 0.665 (0.358-1.235) | 0.197 (1) |
| MDE |  | 0 | 14 | 230 | 0.030 | 0 | 13 | 196 | 0.033 | A | 0.912 (0.289-2.874) | 0.874 (1) |
| EUR | chr22:19964293:C:T  (rs165631; p.Leu203=) | 3 | 342 | 13034 | 0.013 | 1 | 125 | 5191 | 0.012 | T | 1.018 (0.792-1.309) | 0.886 (1) |
| AJ |  | 0 | 45 | 1234 | 0.018 | 0 | 16 | 391 | 0.020 | T | 1.661 (0.639-4.314) | 0.298 (1) |
| AMR |  | 0 | 15 | 427 | 0.018 | 0 | 4 | 150 | 0.013 | T | 0.826 (0.236-2.89) | 0.765 (1) |
| SAS |  | 0 | 12 | 319 | 0.019 | 0 | 6 | 198 | 0.015 | T | 0.85 (0.196-3.679) | 0.828 (1) |
| CAS |  | 0 | 17 | 519 | 0.016 | 0 | 14 | 306 | 0.023 | T | 0.613 (0.248-1.511) | 0.287 (1) |
| MDE |  | 1 | 20 | 230 | 0.048 | 0 | 14 | 196 | 0.036 | T | 0.683 (0.229-2.036) | 0.493 (1) |
| EUR | chr22:19969258:G:A  (rs165599; c.*522G>A) | 6301 | 5523 | 13034 | 0.695 | 2550 | 2145 | 5191 | 0.698 | A | 0.998 (0.937-1.063) | 0.942 (1) |
| AFR |  | 54 | 354 | 929 | 0.249 | 104 | 617 | 1617 | 0.255 | A | 1.051 (0.802-1.377) | 0.718 (1) |
| EAS |  | 631 | 1248 | 2547 | 0.496 | 569 | 1146 | 2345 | 0.492 | A | 1.021 (0.897-1.163) | 0.752 (1) |
| AAC |  | 34 | 135 | 279 | 0.364 | 99 | 365 | 793 | 0.355 | A | 1.037 (0.803-1.340) | 0.778 (1) |
| AJ |  | 453 | 588 | 1234 | 0.395 | 145 | 189 | 391 | 0.387 | A | 1.034 (0.830-1.287) | 0.766 (1) |
| AMR |  | 123 | 204 | 427 | 0.473 | 51 | 77 | 150 | 0.437 | A | 1.375 (0.975-1.94) | 0.069 (1) |
| SAS |  | 92 | 170 | 319 | 0.555 | 70 | 90 | 198 | 0.580 | A | 1.131 (0.762-1.677) | 0.542 (1) |
| CAH |  | 144 | 245 | 494 | 0.539 | 63 | 149 | 283 | 0.486 | A | 1.198 (0.931-1.542) | 0.161 (1) |
| CAS |  | 195 | 245 | 519 | 0.612 | 121 | 139 | 306 | 0.623 | A | 1.007 (0.785-1.291) | 0.958 (1) |
| MDE |  | 74 | 112 | 230 | 0.565 | 63 | 103 | 196 | 0.584 | A | 0.874 (0.579-1.318) | 0.520 (1) |
| EUR | chr22:19969340:G:A  (rs36082074; c.*604G>A) | 9 | 706 | 13034 | 0.028 | 5 | 271 | 5191 | 0.027 | A | 0.933 (0.785-1.109) | 0.431 (1) |
| AJ |  | 6 | 120 | 1234 | 0.053 | 1 | 47 | 391 | 0.063 | A | 0.686 (0.452-1.042) | 0.077 (1) |
| AMR |  | 1 | 13 | 427 | 0.018 | 1 | 9 | 150 | 0.037 | A | 0.46 (0.164-1.291) | 0.14 (1) |
| SAS |  | 1 | 17 | 319 | 0.030 | 1 | 12 | 198 | 0.035 | A | 1.543 (0.547-4.351) | 0.412 (1) |
| CAH |  | 0 | 14 | 494 | 0.014 | 1 | 10 | 283 | 0.021 | A | 0.693 (0.281-1.705) | 0.424 (1) |
| CAS |  | 2 | 28 | 519 | 0.031 | 1 | 15 | 306 | 0.028 | A | 1.032 (0.429-2.481) | 0.944 (1) |
| MDE |  | 1 | 25 | 230 | 0.059 | 0 | 31 | 196 | 0.079 | A | 0.592 (0.275-1.277) | 0.182 (1) |
| EUR | chr22:19969362:T:C  (rs35478083; c.*626T>C) | 20 | 951 | 13034 | 0.038 | 10 | 391 | 5191 | 0.040 | C | 0.968 (0.833-1.126) | 0.677 (1) |
| AFR |  | 6 | 131 | 929 | 0.077 | 9 | 272 | 1617 | 0.090 | C | 1.049 (0.693-1.590) | 0.820 (1) |
| EAS |  | 15 | 331 | 2547 | 0.071 | 11 | 237 | 2345 | 0.055 | C | 0.892 (0.683-1.165) | 0.401 (1) |
| AAC |  | 0 | 39 | 279 | 0.070 | 2 | 127 | 793 | 0.083 | C | 0.624 (0.379-0.028) | 0.064 (1) |
| AJ |  | 0 | 60 | 1234 | 0.024 | 0 | 18 | 391 | 0.023 | C | 1.06 (0.509-2.207) | 0.876 (1) |
| AMR |  | 3 | 36 | 427 | 0.049 | 1 | 14 | 150 | 0.053 | C | 1.822 (0.752-4.414) | 0.184 (1) |
| SAS |  | 2 | 25 | 319 | 0.045 | 0 | 19 | 198 | 0.048 | C | 0.602 (0.239-1.515) | 0.281 (1) |
| CAH |  | 3 | 53 | 494 | 0.060 | 1 | 36 | 283 | 0.067 | C | 0.928 (0.565-1.522) | 0.767 (1) |
| CAS |  | 3 | 53 | 519 | 0.057 | 1 | 37 | 306 | 0.064 | C | 0.808 (0.499-1.307) | 0.385 (1) |
| MDE |  | 0 | 25 | 230 | 0.054 | 0 | 22 | 196 | 0.056 | C | 1.08 (0.461-2.529) | 0.859 (1) |
| EUR | chr22:19969500:C:T  (rs165728; c.*764C>T) | 40 | 1295 | 13034 | 0.053 | 19 | 481 | 5191 | 0.050 | C | 0.984 (0.862-1.124) | 0.817 (1) |
| AFR |  | 0 | 86 | 929 | 0.046 | 7 | 165 | 1617 | 0.056 | C | 0.642 (0.376-1.096) | 0.104 (1) |
| EAS |  | 358 | 1192 | 2547 | 0.384 | 362 | 1069 | 2345 | 0.397 | C | 0.952 (0.831-1.091) | 0.482 (1) |
| AAC |  | 3 | 42 | 279 | 0.086 | 4 | 101 | 793 | 0.036 | C | 0.873 (0.556-1.369) | 0.553 (1) |
| AJ |  | 1 | 93 | 1234 | 0.038 | 2 | 45 | 391 | 0.063 | C | 0.602 (0.373-0.971) | 0.038 (1) |
| AMR |  | 26 | 117 | 427 | 0.198 | 2 | 39 | 150 | 0.143 | C | 1.414 (0.883-2.264) | 0.149 (1) |
| SAS |  | 4 | 76 | 319 | 0.132 | 1 | 44 | 198 | 0.118 | C | 0.913 (0.498-1.674) | 0.769 (1) |
| CAH |  | 15 | 105 | 494 | 0.137 | 7 | 53 | 283 | 0.118 | C | 1.082 (0.752-1.556) | 0.671 (1) |
| CAS |  | 14 | 164 | 519 | 0.186 | 6 | 77 | 306 | 0.146 | C | 1.26 (0.909-1.745) | 0.165 (1) |
| MDE |  | 2 | 21 | 230 | 0.055 | 0 | 13 | 196 | 0.033 | C | 2.475 (1.03-5.949) | 0.043 (1) |
| AFR | chr22:19961219:C:G  (rs11569715; c.-71C>G) | 0 | 68 | 929 | 0.037 | 1 | 109 | 1617 | 0.034 | G | 1.126 (0.561-2.259) | 0.738 (1) |
| AAC |  | 0 | 7 | 279 | 0.013 | 0 | 41 | 793 | 0.026 | G | 0.439 (0.16-1.207) | 0.111 (1) |
| AFR | chr22:19963714:C:T  (rs8192488; p.Ala146=) | 12 | 152 | 929 | 0.096 | 10 | 288 | 1617 | 0.096 | T | 0.916 (0.610-1.375) | 0.672 (1) |
| AAC |  | 1 | 39 | 279 | 0.074 | 2 | 107 | 793 | 0.070 | T | 1.276 (0.801-2.004) | 0.311 (1) |
| CAH |  | 2 | 27 | 494 | 0.031 | 1 | 17 | 283 | 0.034 | T | 0.936 (0.473-1.85) | 0.848 (1) |
| AFR | chr22:19968894:G:A  (rs9332380; c.*158G>A) | 2 | 63 | 929 | 0.036 | 2 | 142 | 1617 | 0.045 | A | 0.882 (0.431-1.806) | 0.732 (1) |
| AAC |  | 0 | 17 | 279 | 0.030 | 0 | 59 | 793 | 0.037 | A | 0.723 (0.347-1.509) | 0.388 (1) |
| AFR | chr22:19969244:T:C  (rs112696498; c.*508T>C) | 6 | 113 | 929 | 0.067 | 5 | 198 | 1617 | 0.064 | C | 1.128 (0.722-1.764) | 0.597 (1) |
| AAC |  | 2 | 34 | 279 | 0.068 | 1 | 61 | 793 | 0.040 | C | 1.538 (0.905-2.612) | 0.111 (1) |
| CAH |  | 1 | 21 | 494 | 0.023 | 1 | 17 | 283 | 0.034 | C | 1.051 (0.529-2.087) | 0.886 (1) |
| AFR | chr22:19969444:C:T  (rs35481270; c.*708C>T) | 0 | 15 | 929 | 0.008 | 1 | 40 | 1617 | 0.013 | T | 1.368 (0.550-3.402) | 0.500 (1) |
| AFR | chr22:19969728:T:C  (rs60537793; c.*992T>C) | 6 | 111 | 929 | 0.066 | 5 | 197 | 1617 | 0.064 | C | 1.080 (0.685-1.701) | 0.742 (1) |
| AAC |  | 2 | 32 | 279 | 0.065 | 1 | 57 | 793 | 0.037 | C | 1.658 (0.968-2.838) | 0.065 (1) |
| CAH |  | 0 | 20 | 494 | 0.020 | 1 | 12 | 283 | 0.025 | C | 1.057 (0.489-2.285) | 0.888 (1) |
| EAS | chr22:19962740:G:T  (rs6267; p.Ala72Ser) | 1 | 118 | 2547 | 0.025 | 0 | 91 | 2345 | 0.021 | T | 0.643 (0.415-0.998) | 0.049 (1) |
| AMR |  | 0 | 30 | 427 | 0.035 | 0 | 3 | 150 | 0.010 | T | 2.863 (0.642-12.774) | 0.168 (1) |
| CAS |  | 0 | 44 | 519 | 0.042 | 1 | 22 | 306 | 0.039 | T | 0.901 (0.508-1.599) | 0.722 (1) |
| CAH |  | 1 | 15 | 494 | 0.017 | 0 | 5 | 283 | 0.009 | T | 0.658 (0.186-2.331) | 0.517 (1) |
| AJ | chr22:19962643:C:T  (rs74745580; c.-34C>T) | 1 | 80 | 1234 | 0.033 | 0 | 13 | 391 | 0.017 | T | 1.911 (0.903-4.046) | 0.09 (1) |
| AJ | chr22:19968753:C:G  (rs368314788; c.*17C>G) | 0 | 47 | 1234 | 0.019 | 0 | 10 | 391 | 0.013 | G | 1.134 (0.497-2.586) | 0.766 (1) |
| AJ | chr22:19968866:C:T  (rs749822167; c.*130C>T) | 0 | 47 | 1234 | 0.019 | 0 | 10 | 391 | 0.013 | T | 1.134 (0.497-2.586) | 0.766 (1) |
| AMR | chr22:19962362:C:T  (rs6268; c.-165C>T) | 3 | 28 | 427 | 0.041 | 0 | 16 | 150 | 0.054 | T | 0.717 (0.338-1.521) | 0.386 (1) |
| EAS |  | 0 | 53 | 2547 | 0.011 | 0 | 51 | 2345 | 0.012 | T | 1.021 (0.52-2.007) | 0.951 (1) |
| SAS | chr22:19969314:G:A  (rs551937723; c.*578G>A) | 0 | 9 | 319 | 0.014 | 0 | 6 | 198 | 0.015 | A | 0.518 (0.116-2.323) | 0.39 (1) |

**Supplementary Table 5.** Gene burden analysis of *COMT* variants.

*Abbreviations: N Var, number of variants.*

| **Cohort** | **Kernel** | **Coding Variants** | | **Potentially Functional** | |
| --- | --- | --- | --- | --- | --- |
|  |  | N Var | *p* | N Var | *p* |
| GP2 AAC | Skat | 4 | 0.415 | 34 | 0.094 |
|  | SkatO | 4 | 0.477 | 34 | 0.176 |
| GP2 AFR | Skat | 8 | 0.957 | 44 | 0.923 |
|  | SkatO | 8 | 1 | 44 | 1 |
| GP2 AJ | Skat | 3 | 0.069 | 28 | 0.219 |
|  | SkatO | 3 | 0.036 | 28 | 0.189 |
| GP2 AMR | Skat | 2 | 0.159 | 26 | 0.207 |
|  | SkatO | 2 | 0.151 | 26 | 0.263 |
| GP2 CAH | Skat | 4 | 0.469 | 33 | 0.797 |
|  | SkatO | 4 | 0.385 | 33 | 0.750 |
| GP2 MDE | Skat | 2 | 0.270 | 16 | 0.0817 |
|  | SkatO | 2 | 0.271 | 16 | 0.164 |
| GP2 CAS | Skat | 2 | 0.723 | 17 | 0.744 |
|  | SkatO | 2 | 0.722 | 17 | 0.165 |
| GP2 EAS | Skat | 8 | 0.604 | 46 | 0.698 |
|  | SkatO | 8 | 0.403 | 46 | 0.520 |
| GP2 EUR | Skat | 18 | 0.020 | 105 | 0.032 |
|  | SkatO | 18 | 0.019 | 105 | 0.037 |
| GP2 SAS | Skat | 4 | 0.170 | 20 | 0.610 |
|  | SkatO | 4 | 0.212 | 20 | 0.596 |
| AMP-PD | Skat | 4 | 0.912 | 46 | 0.071 |
|  | SkatO | 4 | 0.781 | 46 | 0.109 |

**Supplementary Table 6.** *COMT* exonic variants associated with MoCA scores adjusted by age at baseline, sex, education level, and five principal components in GP2 European genotyping data. *Abbreviations: A1, Allele 1, effect allele; BONF, Bonferroni adjusted P-value.*

| **Variant** | **A1** | **BETA (SE)** | ***p***  **(BONF)** |
| --- | --- | --- | --- |
| chr22:19962712:C:T  (rs4633; p.His62=) | C | -0.330 (0.149) | 0.027 (1) |
| chr22:19963748:G:A  (rs4680; p.Val158Met) | G | -0.338 (0.150) | 0.024 (1) |
| chr22:19962429:A:G  (rs6269; c.-98A>G) | G | -0.033 (0.154) | 0.831 (1) |
| chr22:19963684:C:G  (rs4818; p.Leu136=) | G | -0.031 (0.154) | 0.843 (1) |
| chr22:19964281:G:A  (rs769224; p.Pro199=) | A | 0.775 (0.554) | 0.162 (1) |
| chr22:19964293:C:T  (rs165631; p.Leu203=) | T | 0.103 (0.555) | 0.853 (1) |
| chr22:19969258:G:A  (rs165599; c.*522G>A) | G | -0.277 (0.167) | 0.103 (1) |
| chr22:19969340:G:A  (rs36082074; c.*604G>A) | A | 0.519 (0.484) | 0.284 (1) |
| chr22:19969362:T:C  (rs35478083; c.*626T>C) | C | -0.007 (0.409) | 0.987 (1) |
| chr22:19969500:C:T  (rs165728; c.*764C>T) | C | -0.591 (0.354) | 0.095 (1) |

**Supplementary Table 7.** *COMT* exonic variants associated with MOCA scores adjusted by age at baseline, sex, education level, and five principal components in AMP-PD WGS. *Abbreviations: A1: Allele 1, effect allele; BONF, Bonferroni adjusted P-value.*

| **Variant** | **A1** | **BETA (SE)** | ***p* (BONF)** |
| --- | --- | --- | --- |
| chr22:19962429:A:G  (rs6269; c.-98A>G) | G | -0.236 (0.132) | 0.074 (1) |
| chr22:19962712:C:T  (rs4633; p.His62=) | T | 0.183 (0.131) | 0.163 (1) |
| chr22:19963684:C:G  (rs4818; p.Leu136=) | G | -0.218 (0.132) | 0.099 (1) |
| chr22:19963748:G:A  (rs4680; p.Val158Met) | A | 0.161 (0.131) | 0.218 (1) |
| chr22:19964281:G:A  (rs769224; p.Pro199=) | A | 0.203 (0.5496) | 0.712 (1) |
| chr22:19964293:C:T  (rs165631; p.Leu203=) | T | 0.968 (0.680) | 0.155 (1) |
| chr22:19969030:C:G  (rs9332381; c.*294C>G) | G | 0.278 (0.391) | 0.477 (1) |
| chr22:19969258:G:A  (rs165599; c.*522G>A) | A | 0.151 (0.142) | 0.287 (1) |
| chr22:19969340:G:A  (rs36082074; c.*604G>A) | A | 0.201 (0.411) | 0.625 (1) |
| chr22:19969362:T:C  (rs35478083; c.*626T>C) | C | 0.238 (0.394) | 0.546 (1) |
| chr22:19969500:C:T  (rs165728; c.*764C>T) | T | -0.163 (0.300) | 0.586 (1) |

**Supplementary Table 8.** *COMT* exonic variants associated with MDS UPDRS Parts III and IV total scores adjusted by age at baseline, sex, education level, and five principal components in GP2 European genotyping data.

*Abbreviations: A1, Allele 1, effect allele; BONF, Bonferroni adjusted P-value.*

| **MDS-UPDRS Part** | **Variant** | **A1** | **BETA (SE)** | ***p* (BONF)** |
| --- | --- | --- | --- | --- |
| III | chr22:19962429:A:G  (rs6269; c.-98A>G) | G | 0.034 (0.470) | 0.941 (1) |
| IV |  |  | 0.079 (0.267) | 0.768 (1) |
| III | chr22:19962712:C:T  (rs4633; p.His62=) | C | 0.154 (0.461) | 0.738 (1) |
| IV |  |  | 0.125 (0.265) | 0.636 (1) |
| III | chr22:19963684:C:G  (rs4818; p.Leu136=) | G | 0.035 (0.471) | 0.941 (1) |
| IV |  |  | 0.144 (0.268) | 0.591 (1) |
| III | chr22:19963748:G:A  (rs4680; p.Val158Met) | G | 0.152 (0.462) | 0.742 (1) |
| IV |  |  | 0.139 (0.264) | 0.599 (1) |
| III | chr22:19964281:G:A  (rs769224; p.Pro199=) | A | - 0.440 (1.776) | 0.804 (1) |
| IV |  |  | 0.044 (1.02) | 0.966 (1) |
| III | chr22:19964293:C:T  (rs165631; p.Leu203=) | T | 1.05 (1.901) | 0.581 (1) |
| IV |  |  | 1.294 (1.233) | 0.295 (1) |
| III | chr22:19969258:G:A  (rs165599; c.*522G>A) | G | -0.514 (0.518) | 0.321 (1) |
| IV |  |  | 0.020 (0.289) | 0.946 (1) |
| III | chr22:19969340:G:A  (rs36082074; c.*604G>A) | A | -2.335 (1.30) | 0.073 (1) |
| IV |  |  | 0.532 (0.707) | 0.452 (1) |
| III | chr22:19969362:T:C  (rs35478083; c.*626T>C) | C | -0.258 (1.244) | 0.835 (1) |
| IV |  |  | -0.239 (0.669) | 0.722 (1) |
| III | chr22:19969500:C:T  (rs165728; c.*764C>T) | C | -0.769 (1.058) | 0.467 (1) |
| IV |  |  | 0.570 (0.544) | 0.295 (1) |

**Supplementary Table 9.** *COMT* exonic variants associated with MDS UPDRS III and IV scores adjusted by age at baseline, sex, education level and five principal components in AMP-PD WGS. *Abbreviations: A1: Allele 1, effect allele; BONF, Bonferroni adjusted P-value.*

| **MDS-UPDRS Part** | **Variant** | **A1** | **BETA (SE)** | ***p* (BONF)** |
| --- | --- | --- | --- | --- |
| III | chr22:19962429:A:G  (rs6269; c.-98A>G) | G | 0.0112 (0.395) | 0.977 (1) |
| IV |  |  | -0.158 (0.140) | 0.258 (1) |
| III | chr22:19962712:C:T  (rs4633; p.His62=) | T | -0.139 (0.388) | 0.720 (1) |
| IV |  |  | 0.307 (0.138) | 0.026 (1) |
| III | chr22:19963684:C:G  (rs4818; p.Leu136=) | G | -0.053 (0.395) | 0.894 (1) |
| IV |  |  | -0.123 (0.140) | 0.377 (1) |
| III | chr22:19963748:G:A  (rs4680; p.Val158Met) | A | -0.038 (0.388) | 0.921 (1) |
| IV |  |  | 0.298 (0.138) | 0.031 (1) |
| III | chr22:19964281:G:A  (rs769224; p.Pro199=) | A | -1.065 (1.656) | 0.520 (1) |
| IV |  |  | 0.028 (0.584) | 0.962 (1) |
| III | chr22:19964293:C:T  (rs165631; p.Leu203=) | T | 1.872 (1.938) | 0.334 (1) |
| IV |  |  | -0.148 (0.731) | 0.839 (1) |
| III | chr22:19969030:C:G  (rs9332381; c.*294C>G) | G | -2.037 (1.115) | 0.068 (1) |
| IV |  |  | 0.230 (0.389) | 0.555 (1) |
| III | chr22:19969258:G:A  (rs165599; c.*522G>A) | A | -0.327 (0.432) | 0.450 (1) |
| IV |  |  | 0.170 (0.150) | 0.257 (1) |
| III | chr22:19969362:T:C  (rs35478083; c.*626T>C) | C | -2.056 (1.124) | 0.068 (1) |
| IV |  |  | 0.196 (0.392) | 0.617 (1) |
| III | chr22:19969340:G:A  (rs36082074; c.*604G>A) | A | -1.803 (1.135) | 0.112 (1) |
| IV |  |  | -0.579 (0.416) | 0.164 (1) |
| III | chr22:19969500:C:T  (rs165728; c.*764C>T) | T | -0.199 (0.899) | 0.824 (1) |
| IV |  |  | 0.666 (0.320) | 0.038 (1) |

**Supplementary Figure 1.** Locus zoom plot for *COMT* variants versus PD risk. **(A)** Data was obtained from the latest European PD GWAS meta-analysis excluding 23andMe data, consisting of 15,056 PD cases, 18,618 UK Biobank proxy-cases, and 449,056 healthy controls (1). **(B)** Data was obtained from the East Asian PD GWAS, consisting of 6,724 PD cases and 24,851 healthy controls (2). **(C)** Data was obtained from the Latin American PD GWAS, consisting of 807 PD cases and 690 healthy controls (3). **(D)** Data was obtained from the African PD GWAS excluding 23andMe data, consisting of 1,200 PD cases and 2,445 healthy controls (4). **(E)** Data was obtained from the multi-ancestry PD GWAS excluding 23andMe data, consisting of 25,374 PD cases, 18,618 proxy-cases, and 571,138 healthy controls (5). p-values on the log-10 scale are plotted along the horizontal axis with the gene names and size of the flanking region. The most strongly associated SNPs are indicated by a purple diamond and pairwise LD (r2) with these SNPs are indicated by dotted color as described in the legend in the upper right corner. The right vertical axis indicates the regional recombination rate (cM/Mb) which is overlaid in blue. Abbreviations: LD, linkage disequilibrium; PD, Parkinson’s disease; SNP, single nucleotide polymorphism.

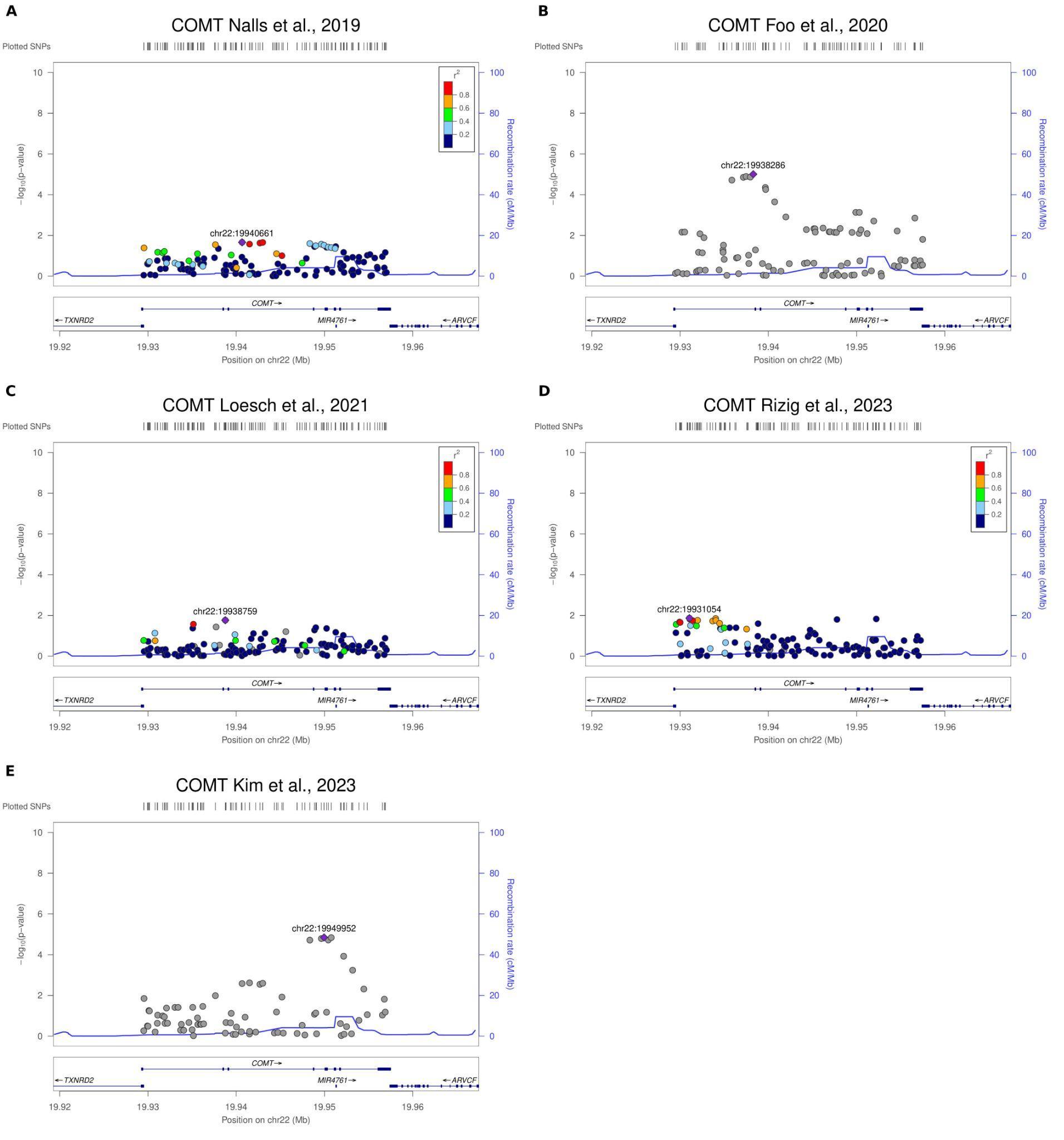

**Supplementary Figure 2.** Levodopa-induced Dyskinesias (LID) survival analysis among 463 PD patients from GP2 European genotyping data.
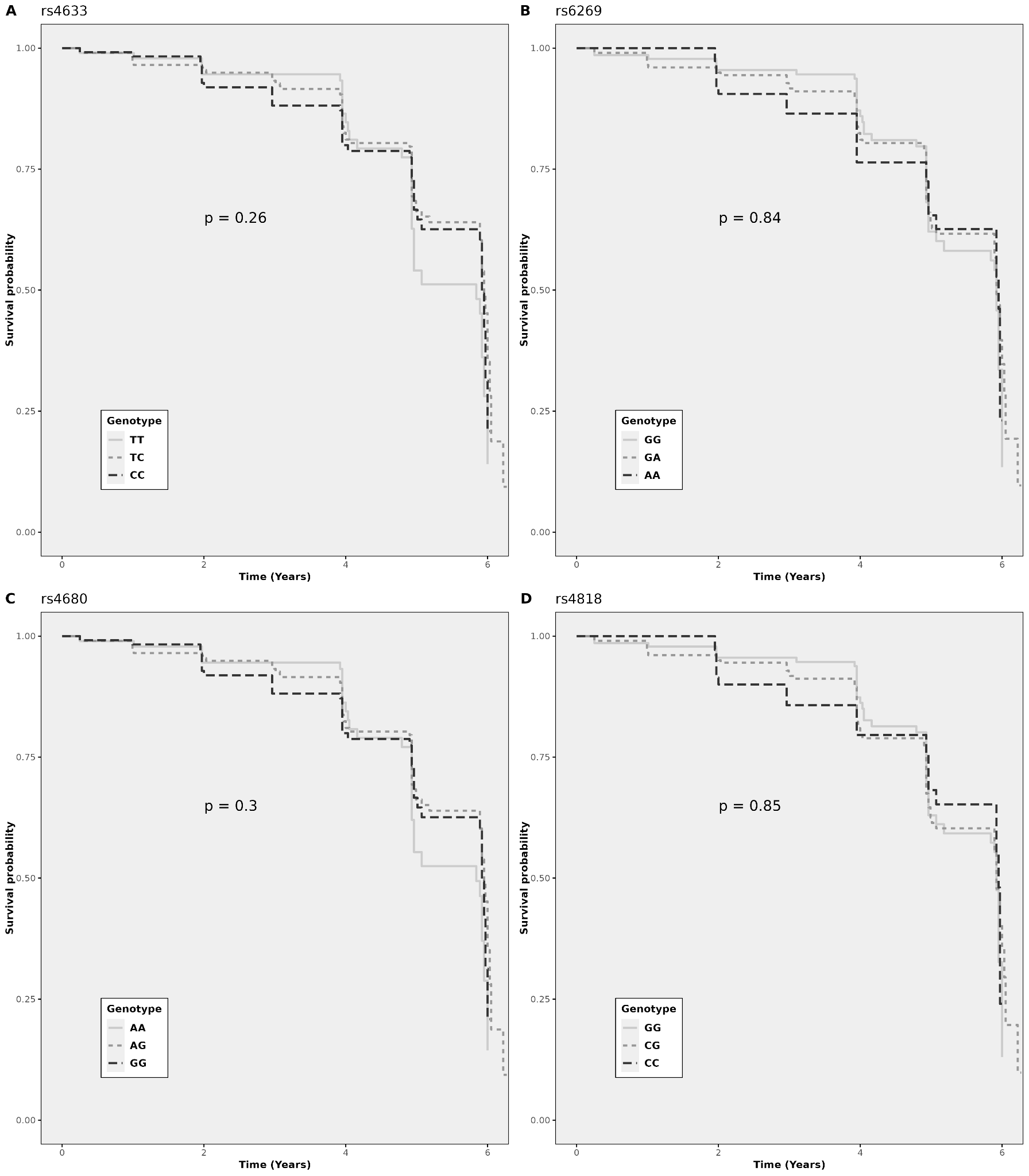
